# eConsent vs. Traditional Consent Among Prospective Biobank Participants: A Randomized Trial

**DOI:** 10.1101/2025.10.30.25339179

**Authors:** Randi L. Vogt-Yerem, Patrick R. Heck, Tamara Gjorgjieva, Rebecca Mestechkin, Daniel Rosica, Anh Huynh, Christopher F. Chabris, H. Lester Kirchner, John Wilbanks, Megan Doerr, Jennifer K. Wagner, Michelle N. Meyer

## Abstract

Electronic consent (eConsent) has the potential to cost-effectively scale research recruitment and improve equity in access to research benefits, but it is unclear whether it achieves comparable informedness compared to traditional consent. In a randomized, controlled, non-inferiority trial (*N*=604; ClinicalTrials.gov ID NCT04131062), we compared a human conversation-based consent process to an eConsent platform similar to that used by NIH’s All of Us Research Program and by studies conducted via Apple ResearchKit. Average comprehension scores of participants randomized to eConsent were non-inferior (*M* = 85.8, SD = 14.7) to those randomized to traditional consent (*M* = 76.5, SD = 22.3; *t*(600.6) = 9.51, *p* < .001). Researchers should give serious consideration to adoption of eConsent as an alternative or supplement to traditional consent. Indeed, the pattern of results we observed suggests that eConsent might yield more informed research enrollment decisions.

## Introduction

Despite widespread consensus on the ethical importance of prospective participants making research enrollment decisions based on adequate comprehension of disclosed information and in light of their own preferences, ensuring that the informed consent process achieves this goal remains a persistent challenge. Research consent forms are often notoriously long and complex, with much content of dubious or at least uneven relevance to individual prospective participants (1–3), many of whom make the arguably rational decision not to read them (4). A systematic review of studies of informed consent for clinical research found that participant understanding of key elements of informed consent, such as the purpose of the treatment or study, the voluntary nature of treatment or research, the ability to withdraw, and the risks and benefits of participation was adequate in only about half of reviewed studies, where the quality of a consent process was deemed “adequate” if more than 80% of the participants in a particular study had a level of understanding (or satisfaction, as applicable) graded in the study’s own highest classification category (5). Although a second systematic review identified several demographic factors associated with poor post-consent comprehension, diverse participants exhibited poor comprehension across the 27 studies reviewed (6).

Traditional research consent processes, in which a member of the study team reviews a paper consent form with a prospective participant, can pose additional challenges (7). Such an approach requires considerable human resources that might be infeasible for large-scale or low-resourced studies (8). In-person consenting in hospitals or clinics risks creating selection biases that could affect the generalizability of the research results and, to the extent that research entails benefits to participants, might create inequities, such as by effectively excluding rural participants or those with transportation challenges (9). The consent “script” that typically serves as the basis for traditional consent conversations is typically one-size-fits-all and relies on the prospective participant to ask to learn more about particular aspects of a study that are confusing or especially relevant to them, which some might be reluctant to do, for a variety of reasons. Human consenters might also make unfounded assumptions about what informational needs or preferences a particular prospective participant has.

Digital approaches to consent (eConsent)—which can be employed in a remote or non-remote fashion (7)—are scalable and personalizable, and so have the potential to address both sets of issues (8,10). They offer additional benefits, such as more seamless integration with electronic health records and other digital platforms, and greater ability to offer participants granular choices (such as opting to permit their data to be shared with some parties but not others, or choosing to receive some research results but not others) and accurately track and implement these choices. And although eConsent enables *user*-driven personalization (e.g., some eConsent platforms permit individuals to learn more about select topics of interest to them, without imposing this extra information on all prospective participants), it can also protect against *human consenter*-driven “personalization” (e.g., consciously or unconsciously tailoring consent discussions to what the consenter perceives the individual wants or needs to know), which can be biased, by providing a common “floor” of information for all.

Despite considerable enthusiasm for eConsent (11,12), scholars have noted the dearth of both high-quality randomized controlled trial (RCT) evidence supporting it, especially in contexts where participants use eConsent to make actual as opposed to hypothetical research enrollment decisions (12) as well as evidence regarding the comparative advantages and disadvantages of eConsent and traditional consent with respect to understanding of information (13). Two systematic reviews of multimedia approaches to consent do suggest, not surprisingly, that simply digitizing the consent process is insufficient to match, much less improve upon, the levels of informedness typically observed following traditional, human-facilitated consent(14,15).

Sage Bionetworks (Sage) has developed an open-source, scalable, participant-navigated eConsent framework that incorporates many elements and approaches recommended in the literature for improving communication and comprehension in clinical and research consent (10,16). In particular, the Sage eConsent framework is designed to present one concisely stated concept per screen, using simple language (14,17). Each screen incorporates an icon that visually reinforces the concept (17,18) (Figure 1). Optional “learn more” hyperlinks on many screens take the participant to an additional page of text with more details, permitting the consent process to efficiently serve people with different informational needs and desires. Finally, formative questions are posed during the consent process that mimic “teach-back” methods and reinforce learning through repeated exposure to the consent material (17–19). In 2015, the Sage eConsent framework was developed by Sage and applied within Apple’s launch of its ResearchKit platform (20), where it has been used to consent participants to decentralized clinical studies conducted via smartphones (21). Since 2018, the U.S. National Institutes of Health *All of Us* Research Program, a large biobank study, has also used the Sage eConsent model, accompanied by brief animated videos. There, the Sage eConsent has been shown to be feasible, with positive indications of informedness about most of the key concepts tested (21,22). However, although promising in terms of participant informedness and satisfaction, as well as scalability and efficiency, it has not been tested against traditional, conversation-based consent.

**Figure 1.**
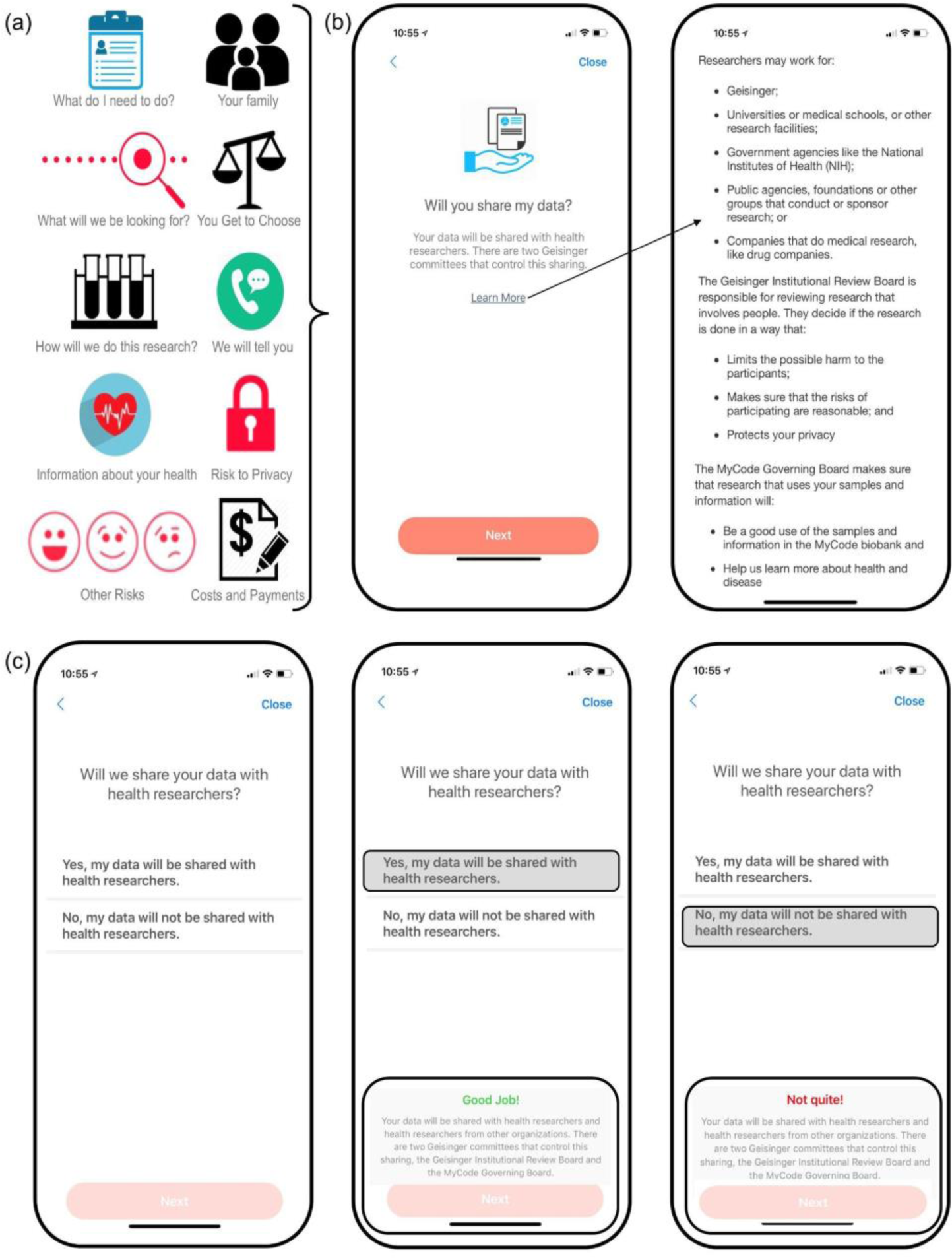
Example eConsent screens *Note.* (a) Example icons used at the top of individual eConsent screens to help convey the main topic of that screen. (b) Example MyCode eConsent screens. On the left, an icon depicting the topic of this screen—data sharing—is displayed first, followed by a concise, two-sentence description of data sharing in MyCode. Here, and in many other (but not all) screens, a “Learn More” link takes users to an optional new screen (right) with additional information about that topic. (c) Example of a “teach-back” question testing users’ knowledge of a recent topic (left) and feedback given to users who get the question right (middle) or wrong (right). Some, but not all, topics are tested by a teach-back question.

### Present Study

In this randomized, controlled trial, we tested whether the Sage eConsent framework (presented using an electronic application on an iPad) was non-inferior to traditional, human-mediated consent in which a consenter reviewed a paper consent with the prospective participant. We pragmatically piggy-backed this trial of eConsent onto the consent process for the MyCode Community Health Initiative (MyCode), the research biobank (23) of Geisinger, a large integrated health system in central and northeastern Pennsylvania. About 3% of MyCode participants receive clinically actionable results (24). All Geisinger patients are eligible to join MyCode and patients are typically approached in primary care clinic waiting rooms, although some consenting occurs in specialty clinics and on inpatient hospital floors. Patients who, after being approached by a consenter, agree to learn more about MyCode then begin the consent process.

In this trial, we randomized patients who agreed to learn about MyCode to either the traditional consent or the eConsent approach (see Figure 2). Those who were able to complete the consent process (which concluded with a decision to enroll in MyCode, not to enroll in MyCode, or to think about it and postpone decision) were asked to complete a survey which asked ten questions that tested their knowledge of MyCode, followed by other questions that measured their perceptions of the consent process and, finally, demographic questions. We conducted preregistered non-inferiority analyses for the outcome of objective comprehension (measured as a “score” on the first 10 survey questions). We conducted exploratory analyses for differences between arms for the remaining outcomes and enrollment decision.

**Figure 2.**
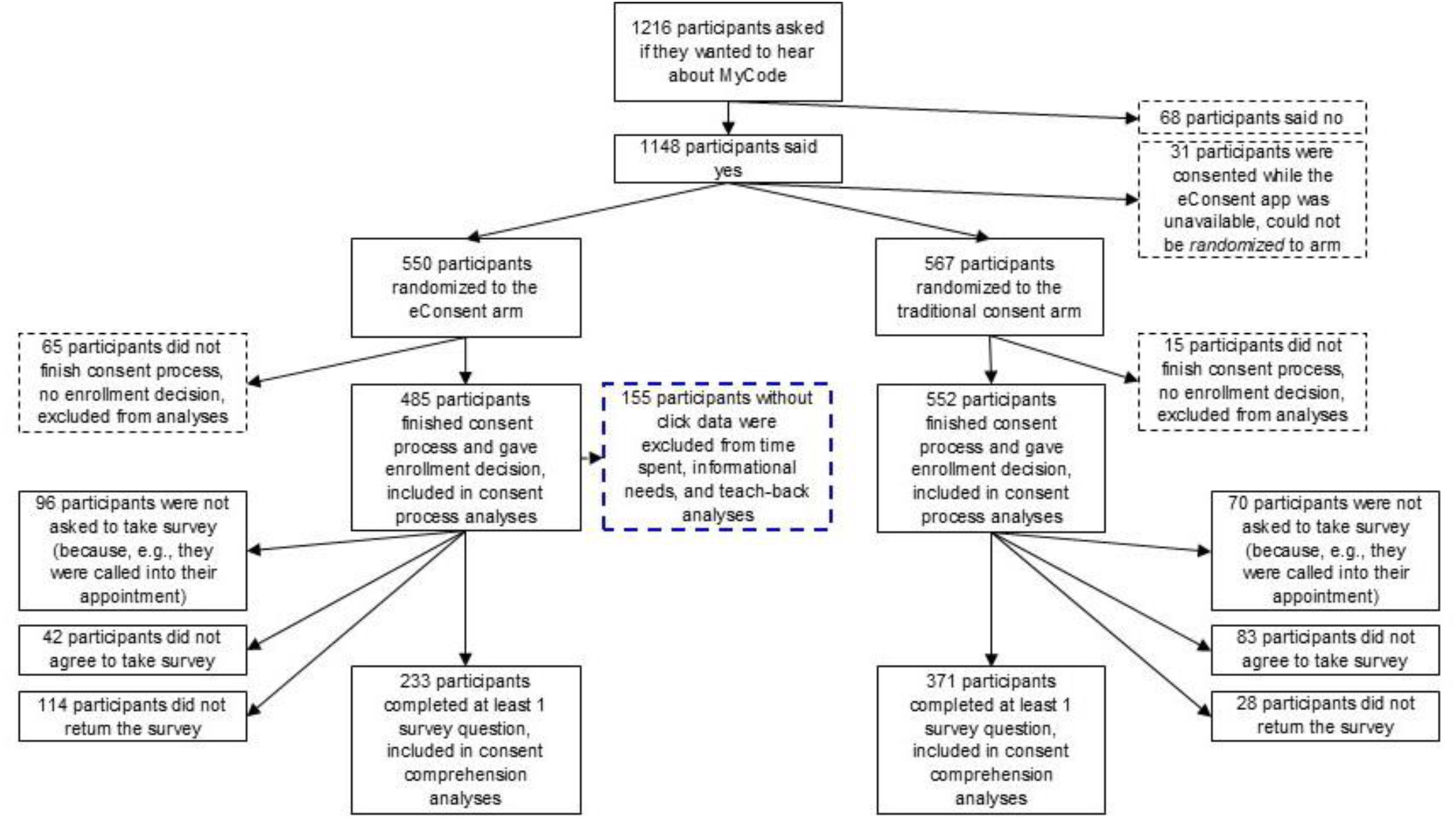
Funnel chart of participants randomized to eConsent arm and traditional consent arm *Note.* The black dashed boxes indicate that these participants are not included in any analyses. The blue dashed box signifies that some data from those participants are missing (because those participants were actually consented through the traditional consent process and not the eConsent process to which they were randomly assigned or because of a bug in the eConsent application on the date of consenting that resulted in data not being recorded) but subsequent data is available so they are excluded from one set of analyses (those analyses specific to metrics recorded in the eConsent app) but included in later analyses (i.e., consent process analyses and consent comprehension analyses).

## Results

### Participant recruitment and baseline characteristics

A total of 1216 Geisinger patients were approached in the usual way about learning more about MyCode; 1148 patients agreed to learn more and 1117 were randomized to either the traditional consent arm or the eConsent arm.^1^ Some participants who were randomized to the eConsent arm were uncomfortable using the iPad and were allowed to switch to the traditional consenting process. However, in our analyses, all participants were analyzed according to intention-to-treat principles (25–27). For instance, if participants were randomized to the eConsent arm but declined to use the iPad, they were consented using the traditional method but included in the eConsent arm for the purpose of our analyses. Using intention-to-treat principles is ideal in pragmatic trials because they allow for deviations from protocol that might occur if the intervention were to be implemented outside of a trial in real-world practice (25–27).

Due to a variety of circumstances (described in Figure 2), only 604 participants (58.2%; 371 participants [67.2%] in the traditional consent arm, 233 participants [48.0%] in the eConsent arm) answered at least one survey question and thus were included in our consent comprehension analyses. Across both conditions, 35.6% of participants who answered at least one survey question answered all survey questions (30.7% of participants in the traditional consent arm and 43.3% of participants in the eConsent arm, *p* = .002).

We found that three out of 15 demographic variables^2^ are unbalanced between arms: race/ethnicity (binarized to non-Hispanic Whites vs. all others), whether the participant reported delaying or not getting medical services in the past year because they could not afford them, and income. There are fewer non-Hispanic White participants, low income participants, and participants who delayed medical services because they could not afford them in the traditional consent arm than in the eConsent arm. We discuss possible reasons for this imbalance in the discussion. See Table 1 for details on participant demographics and characteristics.

**Table 1.**
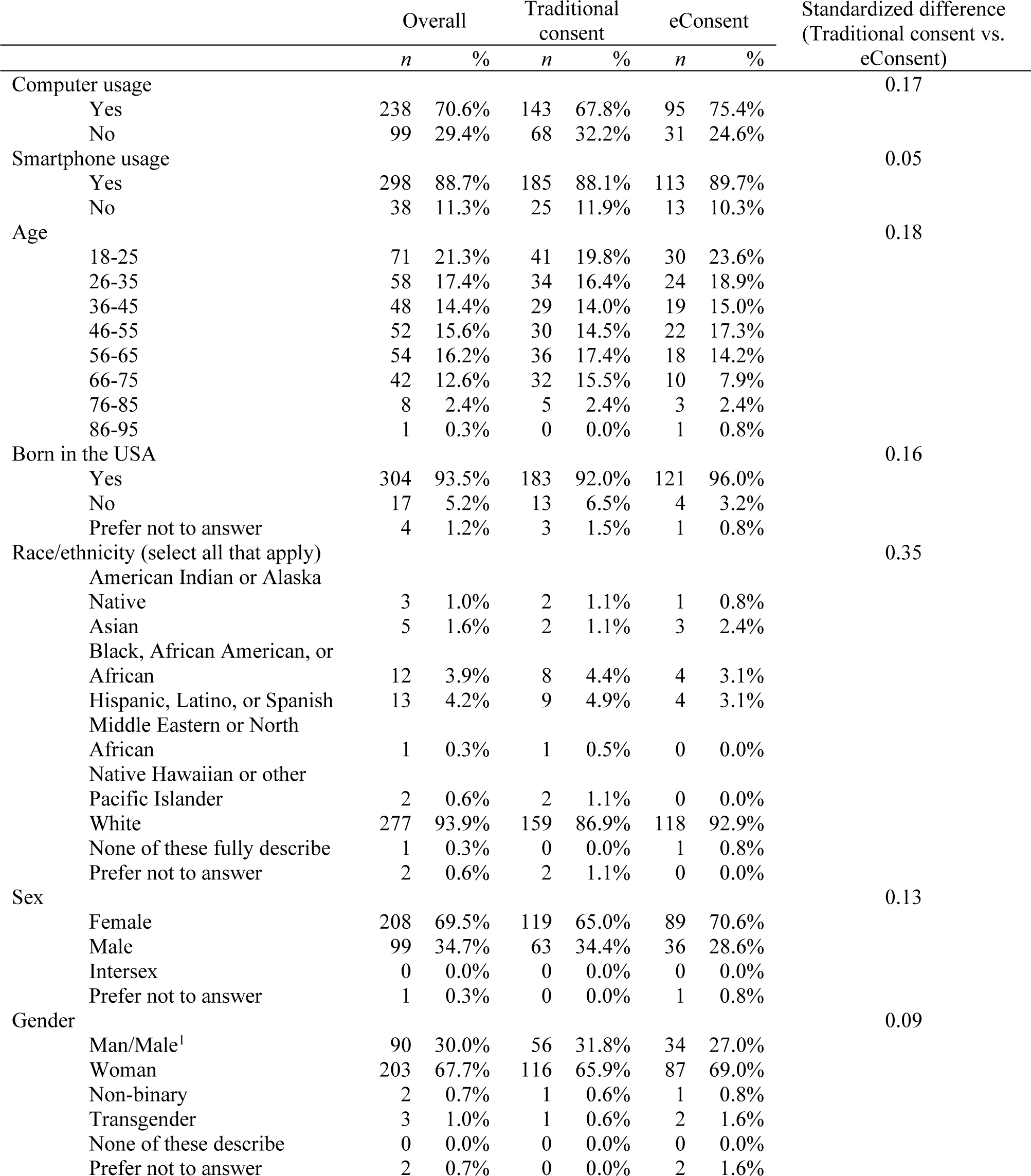
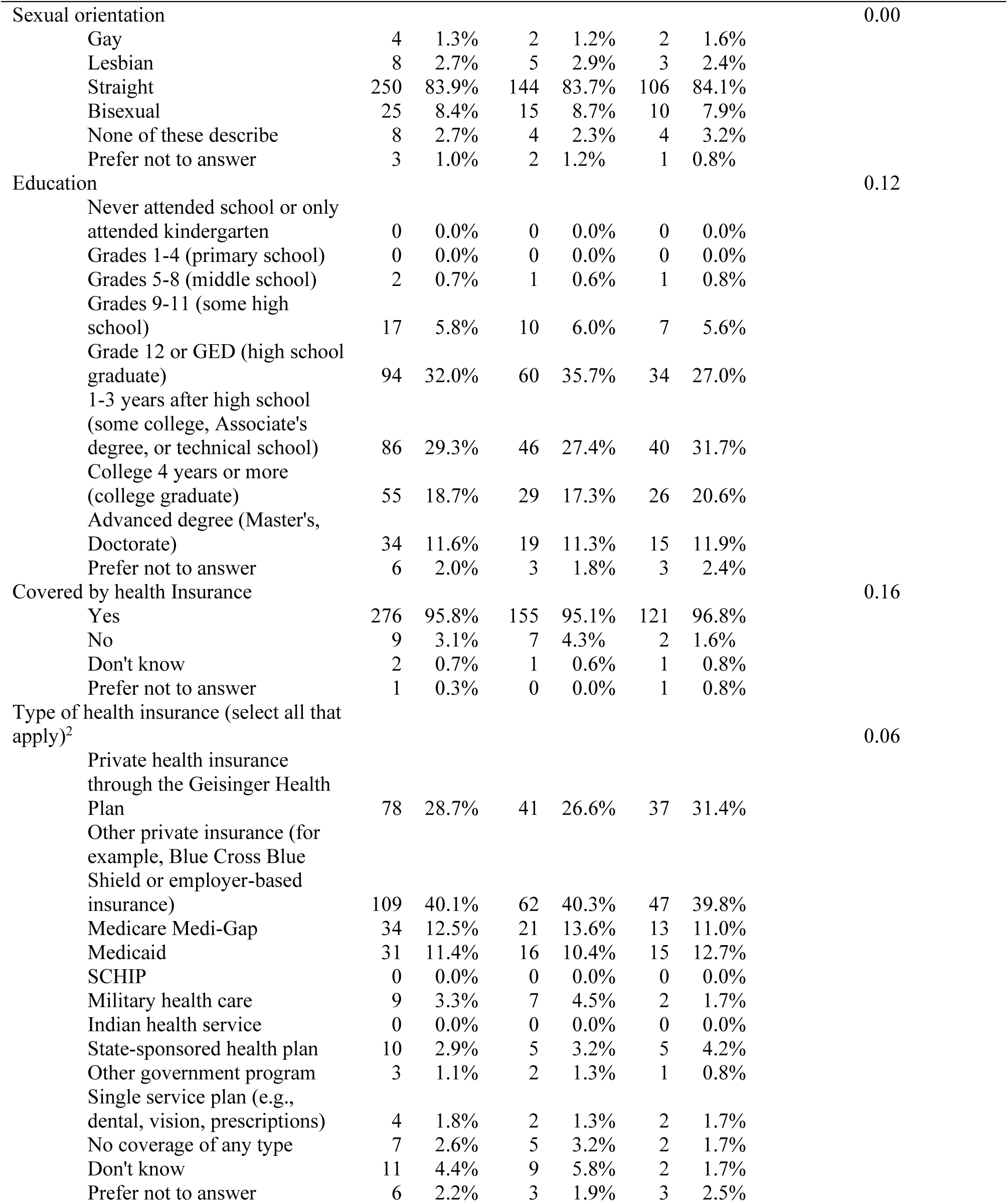
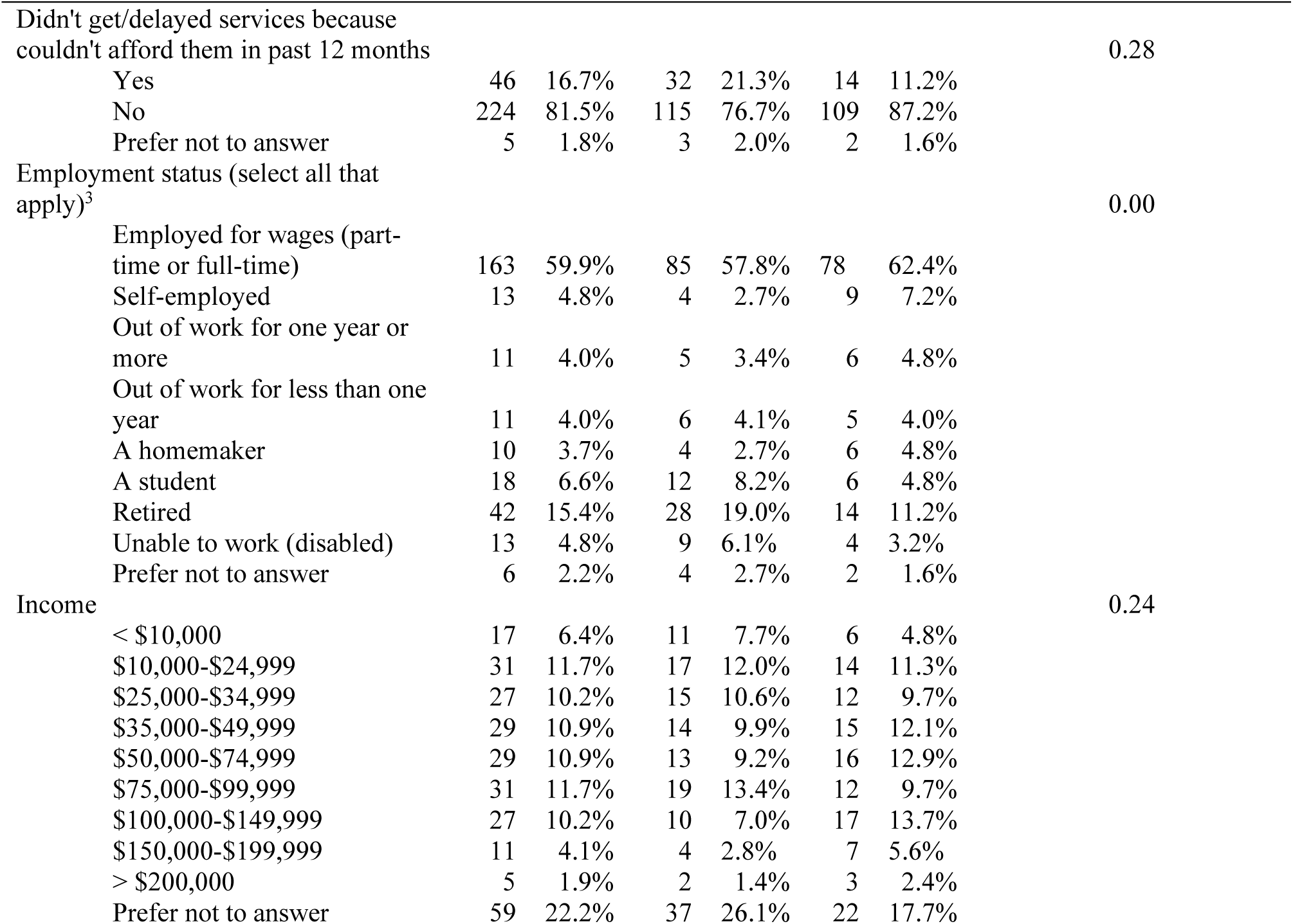
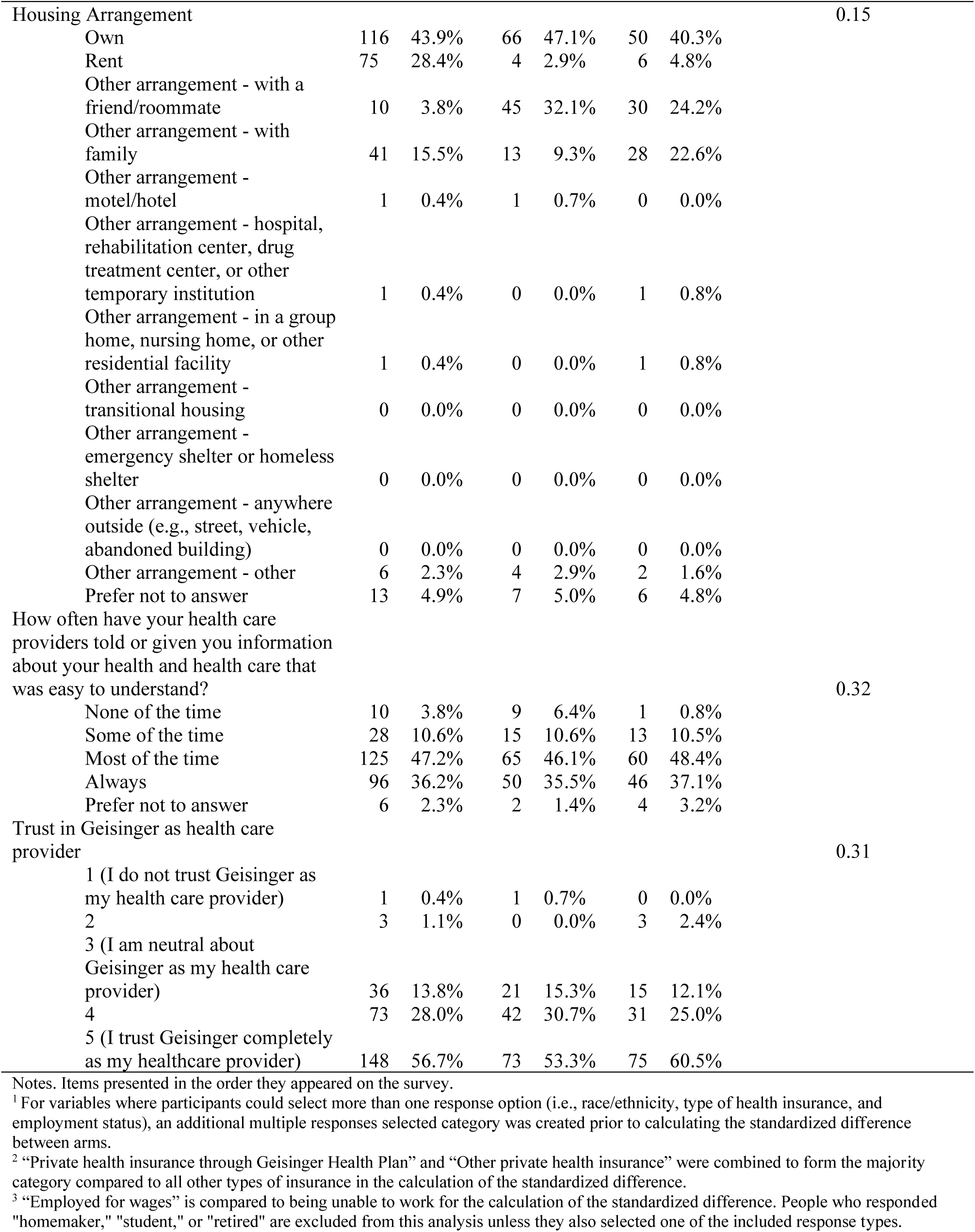
Participant demographics.

### Informedness

Using a policy-meaningful preregistered margin of noninferiority of half of a comprehension question (5 percentage points out of a comprehension score total of 100%), we found that the eConsent process was non-inferior to the traditional consent process on the main outcome of comprehension (eConsent: *M* = 85.8, SD = 14.7, traditional consent: *M* = 76.5, SD = 22.3; *t*(600.6) = 9.51, *p* < .001). However, as noted above, we found that three of our 15 demographic variables were unbalanced between arms. As a robustness check, we repeated this analysis with a subset of our sample that included only participants who reported values for all three unbalanced demographic variables. We excluded those who responded “prefer not to answer.” In this analysis, the eConsent group was non-inferior to the traditional process on comprehension (eConsent: *n* = 101, *M* = 86.0, SD = 13.0, traditional consent: *n* = 99, *M* = 82.3, SD = 14.9; *t*(193.5) = 4.40, *p* < .001). When we further control for those variables in a regression model to adjust for confounding, we similarly found that eConsent was non-inferior to traditional consent (eConsent: *M* = 89.8, SE = 2.1, traditional consent: *M* = 86.5, SE = 2.1; β = .11, *t*(195) = 2.66, *p* = .004).

### Exploratory individual difference and superiority analyses comparing arms

#### Informedness

Using a superiority framework and a conservative ɑ=.005 significance threshold appropriate for exploratory analyses (28) (see Methods), we found that comprehension scores significantly differed between arms in the full dataset (eConsent: *M* = 85.8, SD = 14.7, traditional consent: *M* = 76.5, SD = 22.3; *t*(600.6) = −6.193, *p* < .001). For most (8 of 10) questions, a greater proportion of participants in the eConsent arm selected the correct answer (Table 2). A greater proportion of participants in the traditional consent arm selected the correct answer for two true/false questions: “We will ask you for your social security number as part of this study” and “During this study I will get medical care from MyCode, not from my regular doctor.”

**Table 2.** Comprehension scores, perceived comprehension, and perceived length and difficulty of consent.

|  |  |  |  |  |  |  |  |  |  |
| --- | --- | --- | --- | --- | --- | --- | --- | --- | --- |
|  | Traditional consent |  |  |  | eConsent |  |  |  | *p*<br>(Traditional<br>consent vs.<br>eConsent) |
|  | mean | SD | *n* | % answering<br>correctly | mean | SD | *n* | % answering<br>correctly |  |
| Comprehension quiz |  |  |  |  |  |  |  |  |  |
| Total score | 76.5 | 22.3 |  |  | 85.8 | 14.7 |  |  | <.001 |
| Questions<sup>1</sup> |  |  |  |  |  |  |  |  |  |
| Q1: What is the purpose of MyCode? (4) |  |  | 324 | 87.8 |  |  | 214 | 91.8 | .103 |
| Q2: Who should you call if you have questions about MyCode? (4) |  |  | 279 | 76.4 |  |  | 172 | 79.3 | .426 |
| Q3: Will people who take part in MyCode get information back about their health? (4) |  |  | 174 | 48.3 |  |  | 122 | 52.6 | .313 |
| Q4: What are the possible benefits of taking part in MyCode? (4) |  |  | 347 | 96.9 |  |  | 228 | 98.3 | .282 |
| Q5: While you are taking part in MyCode, your samples and health record... (4) |  |  | 220 | 63.0 |  |  | 199 | 86.9 | <.001 |
| Q6: Which one of the following statements are true? (4) |  |  | 328 | 96.2 |  |  | 221 | 96.5 | .842 |
| Q7: My samples and health record will be given a code. In most cases, this code will be used instead of my name or other information that identifies me. (2) |  |  | 300 | 91.7 |  |  | 223 | 98.2 | <.001 |
| Q8: We will ask you for your social security number as part of this study. (2) |  |  | 307 | 97.5 |  |  | 210 | 92.5 | .012 |
| Q9: During this study, I will get my medical care from MyCode, not from my regular doctor. (2) |  |  | 266 | 86.1 |  |  | 192 | 85.3 | .807 |
| Q10: I have to give a blood or saliva sample as part of MyCode. (2) |  |  | 293 | 91.8 |  |  | 219 | 96.9 | .009 |

|  | Traditional consent |  |  |  | eConsent |  |  |  | *p*<br>(Traditional<br>consent vs.<br>eConsent) |
| --- | --- | --- | --- | --- | --- | --- | --- | --- | --- |
|  | mean | SD | *n* | % | mean | SD | *n* | % |  |
| Perceived comprehension<sup>2</sup> |  |  |  |  |  |  |  |  |  |
| Total sum | 22.8 | 7.3 |  |  | 24.2 | 5.6 |  |  | .018 |
| Questions |  |  |  |  |  |  |  |  |  |
| After learning about MyCode,<br>how well do you think you<br>understand it? | 2.9 | 0.8 |  |  | 2.9 | 0.7 |  |  | .520 |
| I don't understand it all |  |  | 8 | 3.1 |  |  | 8 | 3.7 |  |
| I understand it a little |  |  | 64 | 24.8 |  |  | 49 | 22.7 |  |
| I understand it pretty well |  |  | 128 | 49.6 |  |  | 122 | 56.5 |  |
| I understand it very well |  |  | 58 | 22.5 |  |  | 37 | 17.1 |  |
| After learning about MyCode,<br>how well do you think most<br>people would understand it? | 3.0 | 0.7 |  |  | 2.9 | 0.6 |  |  | .085 |
| They wouldn't understand<br>it at all |  |  | 5 | 2.0 |  |  | 3 | 1.4 |  |
| They would understand a<br>little |  |  | 48 | 19.0 |  |  | 46 | 21.6 |  |
| They would understand it<br>pretty well |  |  | 148 | 58.7 |  |  | 140 | 65.7 |  |
| They would understand it<br>very well |  |  | 51 | 20.2 |  |  | 24 | 11.3 |  |
| Do you know whom you should<br>contact if you have questions or<br>concerns about MyCode? | 3.3 | 0.8 |  |  | 3.0 | 0.9 |  |  | <.001 |
| I have no idea |  |  | 8 | 3.3 |  |  | 13 | 6.2 |  |
| I might know |  |  | 24 | 9.8 |  |  | 32 | 15.2 |  |
| I am pretty sure I know |  |  | 93 | 38.0 |  |  | 94 | 44.5 |  |
| I am confident I know |  |  | 120 | 49.0 |  |  | 72 | 34.1 |  |
| Do you know who will be able to<br>see your MyCode data? | 3.0 | 0.8 |  |  | 3.0 | 0.8 |  |  | .951 |
| I have no idea |  |  | 10 | 4.2 |  |  | 6 | 2.8 |  |
| I might know |  |  | 43 | 18.1 |  |  | 43 | 20.3 |  |
| I am pretty sure I know |  |  | 113 | 47.5 |  |  | 99 | 46.7 |  |
| I am confident I know |  |  | 72 | 30.3 |  |  | 64 | 30.2 |  |
|  | mean | SD | *n* | % | mean | SD | *n* | % |  |
| Do you know what you will be asked to do to take part in MyCode? | 3.3 | 0.8 |  |  | 3.3 | 0.7 |  |  | .697 |
| I have no idea |  |  | 7 | 3.0 |  |  | 4 | 1.9 |  |
| I might know |  |  | 22 | 9.4 |  |  | 21 | 10.0 |  |
| I am pretty sure I know |  |  | 90 | 38.3 |  |  | 91 | 43.1 |  |
| I am confident I know |  |  | 116 | 49.4 |  |  | 95 | 45.0 |  |
| Do you know what the risks are of taking part in MyCode? | 2.9 | 1.1 |  |  | 3.1 | 0.9 |  |  | .012 |
| I have no idea |  |  | 36 | 15.7 |  |  | 11 | 5.2 |  |
| I might know |  |  | 34 | 14.8 |  |  | 37 | 17.5 |  |
| I am pretty sure I know |  |  | 82 | 35.8 |  |  | 82 | 38.9 |  |
| I am confident I know |  |  | 77 | 33.6 |  |  | 81 | 38.4 |  |
| Do you know what information the MyCode researchers will collect? | 3.1 | 0.8 |  |  | 3.0 | 0.9 |  |  | .276 |
| I have no idea |  |  | 11 | 5.0 |  |  | 13 | 6.2 |  |
| I might know |  |  | 31 | 14.0 |  |  | 33 | 15.6 |  |
| I am pretty sure I know |  |  | 97 | 43.9 |  |  | 97 | 46.0 |  |
| I am confident I know |  |  | 82 | 37.1 |  |  | 68 | 32.2 |  |
| Do you know what happens if MyCode researchers find information that may be important to your health? | 3.5 | 0.7 |  |  | 3.4 | 0.8 |  |  | .266 |
| I have no idea |  |  | 4 | 1.8 |  |  | 7 | 3.3 |  |
| I might know |  |  | 11 | 5.0 |  |  | 14 | 6.6 |  |
| I am pretty sure I know |  |  | 72 | 33.0 |  |  | 70 | 33.2 |  |
| I am confident I know |  |  | 131 | 60.1 |  |  | 120 | 56.9 |  |
|  | mean | SD | *n* | % | mean | SD | *n* | % |  |
| Subjective experience<sup>2</sup> |  |  |  |  |  |  |  |  |  |
| Questions |  |  |  |  |  |  |  |  |  |
| How long did the informed<br>consent process feel to you? | 3.5 | 0.6 |  |  | 3.1 | 0.6 |  |  | <.001 |
| Very long |  |  | 2 | 0.9 |  |  | 3 | 1.4 |  |
| Pretty long |  |  | 6 | 2.8 |  |  | 22 | 10.5 |  |
| Pretty quick |  |  | 100 | 46.1 |  |  | 140 | 67.0 |  |
| Very quick |  |  | 109 | 50.2 |  |  | 44 | 21.1 |  |
| How difficult did the informed<br>consent process feel to you? | 3.5 | 0.6 |  |  | 3.3 | 0.5 |  |  | .011 |
| Very difficult |  |  | 2 | 0.9 |  |  | 0 | 0.0 |  |
| Pretty difficult |  |  | 1 | 0.5 |  |  | 6 | 2.9 |  |
| Pretty easy |  |  | 102 | 48.1 |  |  | 125 | 59.8 |  |
| Very easy |  |  | 107 | 50.5 |  |  | 78 | 37.3 |  |

Notes.
<sup>1</sup>Numbers in parentheses following each comprehension question indicate the number of response options for that question.
<sup>2</sup>To calculate the means for perceived comprehension and subjective experience questions are treated as Likert-type variables with 1 corresponding to the first response option and 4 corresponding to the last response option.

When subsetting to only participants who reported values for all three unbalanced demographic variables and then controlling for those variables, comprehension scores are no longer significantly different across arms (subset: eConsent: *M* = 86.0, SD = 13.0, traditional consent: *M* = 82.3, SD = 14.9; *t*(193.5) = −1.875, *p* = .062; subset controlling for covariates: eConsent: *M* = 89.8, SE = 2.1, traditional consent: *M* = 86.5, SE = 2.1; β = .11, *t*(195) = 1.66, *p* = .098).

In Table 3, we report the correlations between comprehension quiz score and all sociodemographic variables collected. Comprehension score is only significantly associated (at ɑ=.005), overall and in each arm separately, with educational attainment (overall: *r* = .29, *p* < .001; traditional consent: *r* = .30, *p* < .001; eConsent: *r* = .25, *p* = .005).^3^ Unsurprisingly, people who have more advanced degrees get higher scores on the comprehension quiz, regardless of which type of consent they experienced. Importantly, there is no difference in the size of the correlation between comprehension score and education between the two arms. In fact, for no demographic variable is the correlation between it and comprehension score significantly different across arms.

**Table 3.** Correlations between comprehension quiz score and individual differences.

|  | Overall |  | Traditional consent |  | eConsent |  | *p*<br>(Traditional consent vs. eConsent) |
| --- | --- | --- | --- | --- | --- | --- | --- |
|  | *r* | *p* | *r* | *p* | *r* | *p* |  |
| Using a computer | .18 | <.001 | .17 | .013 | .18 | .046 | .944 |
| Using a smartphone | .12 | .027 | .16 | .024 | .04 | .656 | .305 |
| Age | -.07 | .177 | -.04 | .586 | -.12 | .172 | .458 |
| Born in the USA | -.02 | .703 | -.05 | .488 | .02 | .800 | .530 |
| Non-Hispanic White | -.07 | .201 | -.15 | .045 | .06 | .532 | .078 |
| Female | .06 | .296 | .06 | .404 | .04 | .674 | .837 |
| Woman | .03 | .567 | .01 | .929 | .07 | .450 | .602 |
| Straight | -.10 | .094 | -.11 | .141 | -.08 | .406 | .745 |
| Education | .29 | <.001 | .30 | <.001 | .25 | .005 | .645 |
| Covered by health insurance | .00 | .972 | .02 | .811 | -.09 | .326 | .370 |
| Private insurance | .04 | .484 | -.10 | .230 | .26 | .005 | .004 |
| Delaying health services | -.01 | .841 | -.07 | .427 | .03 | .755 | .444 |
| Being employed full/part time<sup>1</sup> | .16 | .020 | .14 | .160 | .21 | .039 | .613 |
| Income | .28 | <.001 | .19 | .058 | .37 | <.001 | .162 |
| Owning a home | .08 | .189 | .08 | .381 | .12 | .191 | .724 |
| Understanding health & healthcare info | .08 | .210 | .09 | .276 | .01 | .886 | .526 |
| Trusting Geisinger | -.04 | .518 | -.06 | .461 | -.03 | .728 | .799 |
| Perceived comprehension | .26 | <.001 | .25 | <.001 | .26 | <.001 | .830 |
| Consenting felt quick | .14 | .003 | .24 | <.001 | .14 | .038 | .300 |
| Consenting felt easy | .26 | <.001 | .38 | <.001 | .15 | .032 | .010 |
| Time spent completing consent process<sup>2</sup> | .09 | .024 | .00 | .946 | .02 | .770 | .785 |
| MyCode decision | .13 | .001 | .16 | .002 | .21 | .002 | .567 |
| Teach-back score |  |  |  |  | .26 | <.001 |  |

*Notes.* All categorical variables (e.g., gender) are binarized such that 1 equals the level of the variable listed in the table (e.g., woman) and 0 equals all others (e.g., man, non-binary, transgender, none of these describe me). All ordinal/interval variables (e.g., education) are treated linearly such that each level is one increment higher than the previous level (e.g., advanced degree > 4 years or more of college > 1-3 years of college).
In all correlations, “Don’t know” and “Prefer not to answer” were treated as missing values.
<sup>1</sup> Compared to being unable to work. People who responded “homemaker,” “student,” or “retired” are excluded from this analysis unless they also selected one of the included response types.
<sup>2</sup> For individuals in the eConsent condition who experienced a bug where their time spent on a certain page recorded an impossible (negative) value, an extreme (greater than 7 minutes) value, or an NA when all other pages recorded a time, we replaced that impossible, extreme, or NA value with the mean time spent for that page in order to calculate their mean time spent across the whole consent process.

#### Perceived comprehension

We found that participants’ confidence that they understood MyCode did not significantly differ by arm (eConsent: *M* = 24.2, *SD* = 5.6; traditional consent: *M* = 22.8, *SD* = 7.3; *t*(473.3) = 2.38, *p* = .018; Table 3). This result holds when the question “After learning about MyCode, how well do you think most people would understand it?” is removed from the sum (because it references “most people[’s]” understanding rather than the participant’s own understanding; eConsent: *M* = 21.4, *SD* = 5.2, traditional consent: *M* = 19.9, *SD* = 6.8, *t*(472.9) = 2.66, *p* = .008). In robustness analyses subsetting to only participants who gave responses to the three unbalanced variables, there is similarly no difference between arms (sum with “most people” question included: eConsent: *M* = 25.2, *SD* = 4.4; traditional consent: *M* = 25.5, *SD* = 4.6; *t*(196.4) = 0.54, *p* = .587; sum without “most people” question included: eConsent: *M* = 22.4, *SD* = 4.0; traditional consent: *M* = 22.6, *SD* = 4.2; *t*(196.5) = 0.34, *p* = .736). When controlling for those unbalanced variables, again the arms are not significantly different (sum with “most people” question included: β = −.05, *t*(194) = −0.74, *p* = .460; sum without “most people” question included: β = −.04, *t*(194) = −0.50, *p* = .620).

In both arms, higher perceived comprehension scores modestly predicted higher objective comprehension scores (eConsent: *r* = .26, *p* < .001; traditional consent: *r* = .25, *p* < .001, Table 3).

#### Other perceptions of the consenting process: length and difficulty

Although a majority of participants in both arms felt that the informed consent process was “pretty quick” or “very quick” (eConsent arm: 88.1%, traditional consent arm: 96.3%), those in the eConsent arm reported that it felt significantly slower than those in the traditional arm (full sample: eConsent: *M* = 3.1, *SD* = 0.6; traditional consent: *M* = 3.5, *SD* = 0.6; *t*(423) = 6.49, *p* < .001; subset: *t*(192.4) = 3.96, *p* < .001; subset controlling for covariates: β = −0.28, *t*(192) = −3.96, *p* < .001).

There is no significant difference regarding how difficult the informed consent process felt, with 98.6% of participants in the traditional consent arm and 97.1% of participants in the eConsent arm feeling that the consent process was “pretty easy” or “very easy” (full sample: eConsent: *M* = 3.3, *SD* = 0.5; traditional consent: *M* = 3.5, *SD* = 0.6; *t*(418.3) = 2.56, *p* = .011; subset: *t*(192.6) = 0.42, *p* = .676; subset controlling for covariates: β = −0.03, *t*(192) = −0.36, *p* = .720).

In the traditional consent arm but not the eConsent arm, participants who scored better on the comprehension quiz were more likely to find the consenting process to be quick (traditional consent: *r* = .38, *p* < .001; eConsent: *r* = .15, *p* = .032; Table 3). In both arms, participants who scored better on the comprehension quiz were more likely to report finding the consenting process to be easy (traditional consent: *r* = .40, *p* < .001; eConsent: *r* = .18, *p* = .007).

#### MyCode enrollment decision

Compared to participants randomized to the eConsent arm, those who were randomized to the traditional consent arm were significantly more likely to enroll in MyCode (full sample: traditional consent: 67.4%, eConsent: 53.8%, *χ^2^(1)*= 19.441, *p* < .001; subset: traditional consent: 83.8%, eConsent: 64.4%, *χ^2^(1)*= 8.88, *p* = .003; subset controlling for covariates: β = −1.18, *z* = −3.15, *p* = .002). In both arms, higher objective comprehension scores predicted a greater likelihood of enrolling in MyCode (eConsent: *r* = .21, *p* = .002; traditional consent: *r* = .16, *p* = .002, Table 3).

#### Total time spent in the consenting process

In total, participants completed the eConsent process in an average of 209.9 seconds (3.5 minutes; SD = 141.6 seconds) while participants completed the traditional consent process significantly faster, in an average of 125.1 seconds (2.1 minutes; SD = 51.4 seconds; *t*(417.4) = −10.86, *p* < .001). This pattern holds in both the subset (traditional consent: mean = 118.9 seconds, SD = 147.0 seconds; eConsent: mean = 204.9 seconds, SD = 30.3 seconds; *t*(104) = −5.65, *p* < .001) and when controlling for the unbalanced covariates (β = 0.37, *t*(191) = −5.49, *p* < .001). The amount of time spent in the consenting process does not predict higher comprehension quiz scores in either arm (eConsent: *r* = .02, *p* = .770; traditional consent: *r* = .00, *p* = .946, Table 3).

### Descriptive statistics about eConsent application elements

#### Time spent per page

Participants spent between 4 and 15 seconds on average on each required eConsent screen prior to the “Do you want to join MyCode?” page, on which they spent an average of 42 seconds (Table 4). Other than the “Do you want to join MyCode” page, and not including teach-back questions (discussed below), participants spent the greatest amount of time on the “How will we do this research?” (*M* = 14.8 seconds), “What do I need to do?” (*M* = 14.4 seconds), and “Risk to privacy” (*M* = 7.9 seconds) pages.

**Table 4.** Means, SDs, and percentages for metrics captured by the eConsent application.

|  | Time spent per page (seconds) |  | Correct answer (teach-back questions) |  |
| --- | --- | --- | --- | --- |
|  | mean | SD | *n* | % |
| Page title |  |  |  |  |
| Informed consent<sup>1</sup> | 6.3 | 6.5 |  |  |
| Check your understanding<sup>1</sup> | 5.5 | 5.6 |  |  |
| What is MyCode?<sup>1</sup> | 5.5 | 5.8 |  |  |
| What is the goal of MyCode? (teach-back question) | 14.9 | 14.3 | 243 | 73.6 |
| How will we do this research? | 14.8 | 18.6 |  |  |
| What will we be looking for?<sup>1</sup> | 6.7 | 7.2 |  |  |
| What to do I need to do?<sup>1</sup> | 14.4 | 13.8 |  |  |
| What will we study in MyCode? (teach-back question) | 9.5 | 7.8 | 323 | 99.7 |
| Will you share my data?<sup>1</sup> | 7.4 | 9.2 |  |  |
| Will we share your data with health researchers? (teach-back question) | 8.4 | 6.9 | 309 | 95.4 |
| Information about your health<sup>1</sup> | 6.3 | 8.0 |  |  |
| We will tell you | 5.7 | 7.1 |  |  |
| Your family | 7.8 | 8.7 |  |  |
| Will we tell you if researchers find information that could be important to your health? (teach-back question) | 8.6 | 7.2 | 320 | 99.1 |
| Risk to privacy<sup>1</sup> | 7.9 | 11.7 |  |  |
| Other risks<sup>1</sup> | 5.8 | 8.7 |  |  |
| What is the main risk of taking part in MyCode? (teach-back question) | 9.8 | 9.9 | 308 | 95.4 |
| Potential benefits<sup>1</sup> | 5.0 | 8.1 |  |  |
| Costs and payments | 3.6 | 6.0 |  |  |
| How long will MyCode last? | 5.2 | 4.2 |  |  |
| You get to choose<sup>1</sup> | 4.2 | 4.8 |  |  |
| If I join, can I withdraw ("quit")?<sup>1</sup> (teach-back question) | 5.8 | 4.7 | 322 | 99.7 |
| Next step | 7.1 | 17.7 |  |  |
| Do you want to join MyCode?<sup>1</sup> | 41.9 | 80.9 |  |  |
| Mean teachback score | 90.5 | 20.1 |  |  |

Notes. <sup>1</sup>Due to an eConsent application bug, for some individuals (no more than four per page), the time spent recorded for this page was either impossible (a negative number), extreme (greater than 7 minutes which was at least two times the next closest response time), or recorded as "NA" (despite having time recorded for subsequent pages, i.e., not a dropout). The means recorded in this table are calculated without those values included. This error only affected 19 participants in total, with only one instance of this bug per participant (except for one participant who had two "NA"s).

#### Informational needs

Very few participants chose to “learn more” in the eConsent arm (Table 5). Only 53 participants (16.1% of all participants with eConsent click data) chose to “learn more” about at least one topic. The pages with the greatest number of clicks were for “Risk to privacy” (*n* = 21) and “Will you share my data?” (*n* = 19).

**Table 5.**
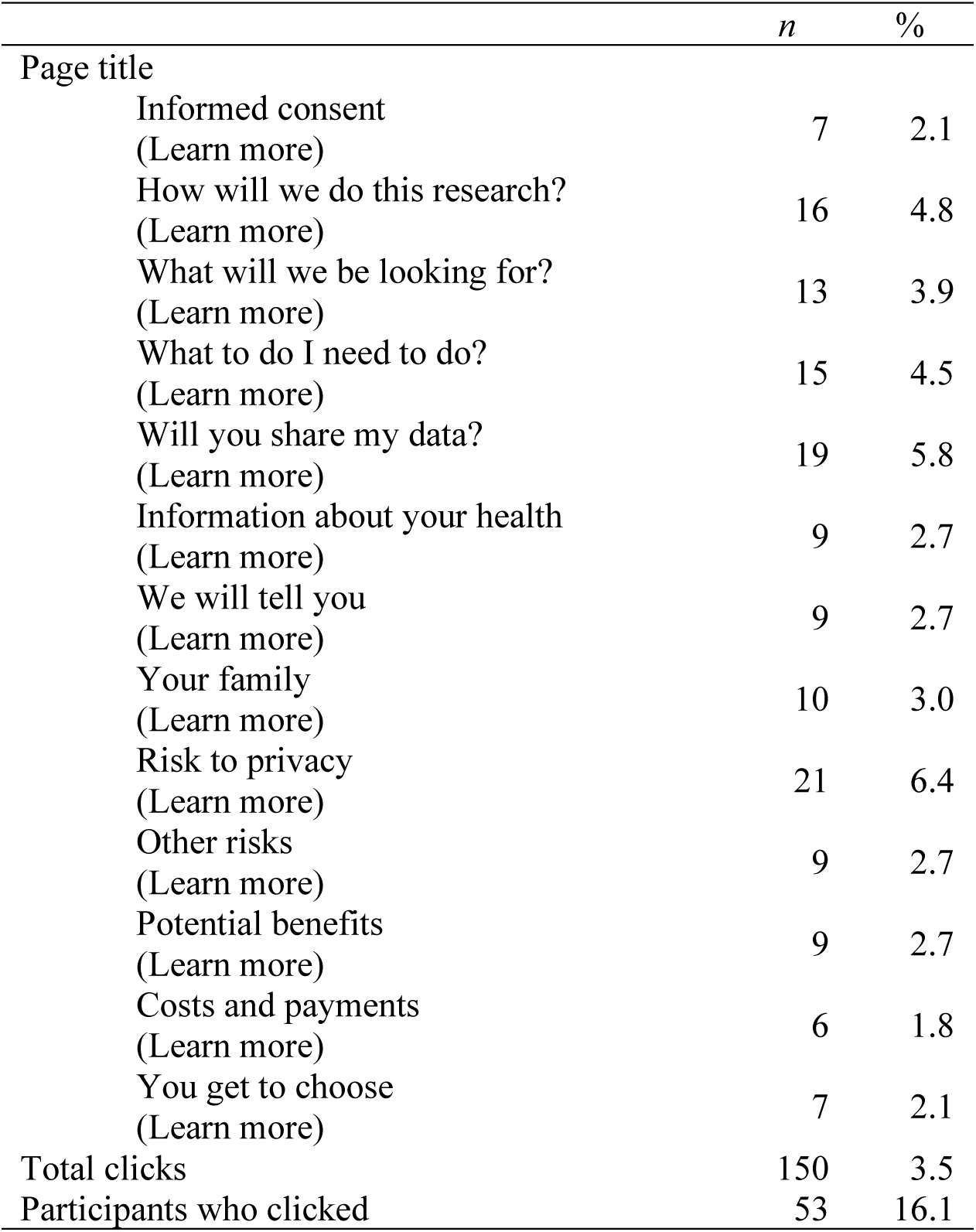
Number of clicks and percentage of participants who clicked on the learn more pages as captured by the eConsent application.

#### Teach-back questions

Overall, participants who experienced the eConsent process performed well on the “teach-back” questions (*M* = 90.5, SD = 20.1). As distinct from the comprehension questions all participants received in the post-consent survey, these questions are administered during the eConsent process and are designed to reinforce participants’ knowledge of recently-explained topics (see Fig. 1(c)). Out of 14 eConsent screens describing an aspect of MyCode, six were reinforced via teach-back questions. Most (67.38%) participants answered all six questions correctly and 24.4% answered five out of six questions correctly. More than 95% of participants correctly answered questions 2-6 (Table 5). Comparatively fewer (73.6%) participants correctly answered question 1 (Q: “What is the goal of MyCode?” A: “To provide healthcare” (incorrect), “To make discoveries about health” (correct)). Five of the six teach-back questions corresponded to topics tested by five survey comprehension quiz questions (with similar but not identical wording), while one teach-back question was similar in topic to two of the comprehension quiz questions. Unsurprisingly, higher teach-back scores were associated with higher comprehension quiz scores overall (*r* = .26, *p* < .001).

## Discussion

These results show that the eConsent process is non-inferior to the traditional consent process in patients’ understanding of the research biobank. Indeed, while we preregistered a non-inferiority trial of eConsent compared to traditional consent with respect to the outcome of participants’ informedness, the directional pattern of comprehension quiz results suggests that the eConsent platform we tested might be superior in producing informedness. The mean comprehension scores in the full sample hint that the participants in the eConsent arm understood the research biobank to which they were consenting *better* than those in the traditional consent arm (although the results in the subset and when controlling for the unbalanced variables in the subset do not reach significance at our conservative ɑ=.005 significance threshold appropriate for exploratory analyses). Our results are consistent with recent randomized, controlled trials that found that other forms of eConsent are either non-inferior or superior to traditional, paper-based consent in terms of informedness (29–32).

We measured participants’ perception of how easy or difficult they felt the consent process was, and found no significant difference between arms. Nor did we find any significant difference in participants’ post-consent confidence that they understood MyCode between arms. While perceived comprehension and objective comprehension were significantly correlated, it was a small correlation, suggesting that some participants overestimated their understanding relative to their actual comprehension. Practitioners and researchers should consider this disconnect between self-reported and objective comprehension when implementing these consenting processes.

However, we did observe some significant differences between arms. Participants randomized to the eConsent process were significantly less likely to enroll in the biobank and to report that the process felt pretty or very quick; the eConsent process was also in fact longer. However, the traditional MyCode consent process is fairly quick; the eConsent platform we tested might be non-inferior in terms of time to other, longer traditional consent processes. Moreover, although the eConsent took on average nearly twice as long as the MyCode traditional consent in relative terms, in absolute terms, it took only 1.4 minutes longer. Finally, although the particular eConsent process we trialed both feels—and objectively is—slightly longer than the particular traditional consent process it was compared to, consent is a rare case where a certain degree of intentional friction might actually be a good thing, enabling more deliberate reflection prior to decision-making. A slightly longer consent process might be an acceptable trade-off in exchange for scalability or other eConsent benefits.

A key strength of the current work is its pragmatic nature: participants made real—not hypothetical—decisions about enrolling in a research biobank. Health system employees continued their usual role as consenters and prospective biobank participants participated in whichever consent process they were randomly assigned to, unaware that both approved consent methods were under comparative evaluation. This minimized the likelihood that patients behaved differently than they would outside of a study, such as paying closer attention in an attempt to help researchers demonstrate a perceived hypothesis about consent (33). However, we only compared one particular form of eConsent to one particular form of conversation-based consent. Additional research is needed to determine whether our results generalize to other instances of traditional consent and eConsent.

The pragmatic nature of the trial also contributed to the attrition that we observed from random assignment to final measured outcome. While the length of the eConsent process might have exacerbated how many participants dropped out before completing the process and providing outcome data, losing prospective MyCode participants because they were called into the exam room is a known issue in this research recruitment process. Nevertheless, if participants who dropped out were different in outcome-relevant ways than those who were able to see the process through to the end of the survey, the attrition we observed might limit the generalizability of our findings to settings in which similar attrition is not common (e.g., research recruitment in clinics with longer wait times or outside of clinical spaces).

A related limitation of this study is the imbalance, among those who did reach the end of the survey, of some demographic variables between trial arms, such that the sample of eConsent participants were more likely to report being White and higher-income. Various aspects of the design of the study protocol might have contributed to these differences. First, we collected demographic data not only after the consent process concluded but also at the very end of the post-consent survey (following all the comprehension quiz questions, the perceived comprehension questions, and questions about their subjective experience of the consenting process). Most patients participated in this trial in a clinic waiting room; the consent and/or survey process generally stopped whenever they were called back for their appointment. It is possible that the out-of-balance demographics are correlated, for a variety of reasons, with taking longer on the consent process and/or the prior survey questions. For instance, some subgroups might be more skeptical of research and review the consent text more closely, they might generally take more time reviewing text, or they might be less familiar with iPads. In these cases, these groups might have been evenly balanced at randomization and throughout the consent intervention and outcome data collection and merely appear unbalanced due to the lateness of when the demographic data were collected. Second, and relatedly, the intervention itself differed in how much time it took participants to complete (see below), leading to differential attrition between arms after random assignment. The eConsent arm took longer, which meant that fewer people in that arm completed the consent process and even fewer reached the last section of the survey where demographic data were collected. Therefore, whether the imbalances we see in these variables reflect the balance or imbalance of the initial random assignment to consent arm is unknown. Finally, across both arms, 22.2% of participants who answered the income question responded that they preferred not to report their income. This makes it more difficult to determine whether there was true imbalance, even among those who reached the end of the survey, for the income variable.

This pragmatic trial was also not designed to determine *which* elements of the eConsent we investigated—e.g., icons, teach-back questions, limiting each screen to one or two simple sentences conveying a single concept, or relegating details about each concept to optional “learn more” screens—were most helpful in ensuring informedness, whether individually or in aggregate, or why. For instance, it could be that teach-back questions during the eConsent process help participants reinforce and retain what they learn, or it could be that participants who know that their knowledge will be tested via teach-back questions simply pay closer attention during that kind of consent process in general, and holding participants accountable in this way in turn drives improved post-consent informedness. Future research should explore such questions.

The “learn more” information architecture might be effective in personalizing the consent experience: it gives information-seeking individuals opportunities to explore further, while allowing others to stay focused on the most critical pieces of information. However, we observed that few participants in the eConsent arm opted to access this additional content. Therefore, eConsent designers should be careful not to place critical information behind these optional layers—and it is not always obvious, even to ethicists, what information about a study is critical for prospective participants to know (34).

## Methods

### Procedure

Each day, as per usual practice, MyCode consenters received a list of patients scheduled for appointments in selected clinics across the health system’s then two-state catchment area, who were eligible to be approached about MyCode. Any patient who had not previously enrolled in, or declined to enroll in, MyCode was considered eligible. Also as per usual practice, eligible patients were asked if they would like to hear about Geisinger’s MyCode Community Health Initiative—a precision medicine project that collects and stores blood and other samples for use (along with electronic health record data) in genetic research and also screens participants’ whole exomes for clinically actionable variants and returns this information to patients and their providers for variant-appropriate care. If the patient agreed to hear about MyCode, they were randomized to either the traditional consent arm or the eConsent arm based on the time their conversation began. Patients whose conversations began on an odd minute (e.g., 11:47am) according to the consenter’s clock were randomized to the eConsent arm, where the consenting process was initiated on an iPad. Patients whose conversations began on an even minute (e.g., 11:48am) according to the consenter’s clock were randomized to the traditional consent arm, in which the consenting process was initiated on paper.

In the traditional consent arm, the consenter went through a standard script that the MyCode team developed from the written consent form that highlighted the most important aspects of MyCode, including return of actionable results to participants and their primary care physicians, genetic privacy, and data sharing for research purposes. At the end of the script, the consenter invited and answered questions from the patient. Next, the consenter handed the patient the 7-page written consent form and offered them a chance to review it. Finally, the consenter asked the patient whether they wished to enroll in MyCode and documented their response—Yes, No, or Thinking (i.e., the patient wanted more time to consider)—in the patient’s electronic health record.

The eConsent arm consisted of an entirely digital version of the informed consent process using an application which interactively walked participants through MyCode. The application includes optional “learn more” links which participants can click if they want more information about the topic on the main page. In addition, there are six teach-back questions, each with only two possible answers. Participants were given feedback (e.g., “Good Job! [explanation]” or “Not quite! [explanation]”) letting them know if they had answered the teach-back question correctly. As in the traditional consent arm, in the eConsent arm, the consenter recorded whether the patient asked any questions about MyCode, when a patient declined to hear about MyCode, and when the consent process was interrupted.

In each arm, the length of the consenting process was timed. In the traditional consent arm, this was done using a stopwatch, which began the moment the participant was introduced to MyCode and concluded when a participant either finished or was interrupted during (e.g., because the patient is called back to the examination room) the consenting process. In the eConsent arm, this was measured internally by the application. The application measured how long (in seconds) each person spent on each page.

After the consenting process concluded, all participants were asked to complete an anonymous survey which included a question about their decision to join MyCode, a 10-question comprehension quiz, a set of eight questions probing their perceived comprehension of MyCode, two questions about their subjective experience of the consenting process, a set of 16 demographic questions, and one item asking them to rate their trust in Geisinger as a healthcare provider. Survey questions are closed-end (true/false, multiple choice, Likert scale) and based on the Quality of Informed Consent and *All of Us* participant-provided information surveys.

Participants in the traditional consent arm completed this survey with pen and paper. Participants in the eConsent arm completed this survey on the iPad using Qualtrics. Overall, of participants who completed the entire consent process, 20.7% of participants completed every question in the survey: 20.8% in the eConsent sample and 20.7% in the traditional consent sample. Data were collected between November 2019 and May 2022 (with relatively few participants recruited in 2020 due to the Covid-19 pandemic).

### Sample size calculations

The study sample size was powered on the primary outcome of total comprehension score. Calculations were based on preliminary data on 101 individuals using the paper consent for enrollment into Geisinger’s MyCode biobank. In our work, the estimated standard deviation (SD) was 1.17. In addition, we assumed 99% power and a significance level of 2.5%. Lastly, the non-inferiority margin was set a priori at 0.5 question on the 10-question comprehension quiz (5 points on a 100-point scale). Therefore, the study was required to recruit 204 participants per group to achieve 99% power and 2.5% significance level. Due to an error in our registered power analysis, we overrecruited participants.^4^

### Data analysis

Prior to conducting the primary analyses, we calculated the standardized difference (*d*) for each demographic variable to assess the differences between arms (35). A *d* > 0.2 was considered to be unbalanced. In this calculation, any responses of “prefer not to answer” or “don’t know” were considered missing data. All categorical variables (e.g., race/ethnicity) were binarized such that 1 corresponds to the majority category (e.g., non-Hispanic White) and 0 corresponds to all other categories (e.g., American Indian or Alaska Native, Asian, Black, African American, or African, etc.). All ordinal/interval variables (e.g., income) are treated as continuous, numeric variables (e.g., “<$10,000” = 1, “$10,000-$24,999” = 2, etc.). Answers to the last two questions of the survey (“How often have your health care providers told or given you information about your health and health care that was easy to understand?” and “On a 1-to-5 scale, how would you rate your trust in Geisinger as your health care provider?”) could plausibly have been influenced by the consent intervention. Therefore, we did not calculate the standardized difference between arms for those variables.

In our primary analysis (informedness), we tested for non-inferiority of eConsent vs. traditional consent by calculating the group difference on comprehension total score and the corresponding 1-sided 97.5% confidence limit. Non-inferiority was demonstrated when the 1-sided 97.5% confidence limit did not cross the non-inferiority limit of 0.5 questions on the 10-question comprehension quiz (5 points on a 100-point scale). Because we found three variables to be unbalanced, another sensitivity analysis that adjusted for the imbalance was performed. This was accomplished using a linear regression model regressing comprehensive total score on group assignment and the unbalanced baseline variables. The regression model was then used to estimate adjusted means and the adjusted difference in comprehension between the groups along with 1-sided 97.5% CI (36).

The other outcomes (perceived comprehension, subjective experience, enrollment decision, and total time spent) are of secondary interest and are analyzed on an exploratory basis in a superiority framework using a significance threshold of α=.005 (28). Because it is not obvious whether the value of the eConsent process is demonstrated by being non-inferior or superior to the traditional consent process on these secondary outcomes (i.e., is it better for participants to take more time or less time to complete the eConsent process compared to the traditional consent process?), we discuss only whether there are or are not differences between arms for these variables.

We report descriptive statistics (means, SDs, Ns, and percentages) for the overall comprehension quiz score, the correct answer for each individual quiz question, the overall perceived comprehension sum, and the responses for each individual perceived comprehension question in each consenting arm as well as the *t*- (or χ^2^-) statistic comparing arms. For the eConsent arm only, we report descriptive statistics (means, SDs, Ns, and percentages) for time spent on each application screen, number of clicks on the “Learn More” screens, and responses to the teach-back questions. We also report descriptive statistics (Ns and percentages) for all responses to the socio-demographic survey items across the full sample. To explore the associations between quiz score and the socio-demographic items, we reported correlations.

In preparing the data for analysis, a few issues were identified and, when possible, resolved. In total, these issues affected 16 participant IDs, 1.4% of the entire data set, and thus are unlikely to affect any results.

- In six cases, the incorrect participant ID was entered into Qualtrics. Those IDs were corrected by matching the date automatically generated by Qualtrics to the date associated with the ID on the consenter tracking sheet.
- In two cases, the randomization column of the consenter tracking sheet said the participant was in the traditional consent arm. However, the tracking sheet also indicates that the participants declined using the iPad suggesting that they were actually initially assigned to the eConsent arm and switched to the traditional consent arm.
- In two separate cases, the tracking sheet indicated that the participant was in the traditional consent arm despite there being some eConsent data logged for those participant IDs. In all four of these cases, the randomized arm was manually changed by the authors from traditional consent to eConsent based on the above described evidence.
- In three cases, there were two physical paper surveys with the same participant ID. Using context clues (such as the date the surveys were uploaded and the participant’s MyCode decision as recorded by the consenter and as answered on the survey), we identified one survey as matching the participant ID on the consenter tracking sheet. The correct participant ID of the second survey could not be identified so that survey was not matched to the tracking sheet and is not included in any analyses.
- In two cases, there were two entries in the eConsent data file with the same participant ID. By matching dates, we identified one entry as matching the participant ID on the consenter tracking sheet. In these cases, we rename the second instance of the participant ID and retain that data as we know that that person must have been assigned to the eConsent arm.
- In one case, we have a physical paper survey that does not match any participant ID on the consenter tracking sheet. Because we cannot identify the condition to which this participant was assigned, this data is not included in any of our analyses.

## Data Availability

Access to the de-identified data, R code, and survey materials is available on the project's OSF page and will be released upon final publication of this paper.

## ETHICS

On May 25, 2018, the Geisinger Institutional Review Board (IRB) approved an amendment to the MyCode Community Health Initiative protocol (IRB# 2006-0258) allowing the eConsent that was evaluated in this study to be used as one of three methods of obtaining consent for MyCode (along with the traditional consent process described here and a third method in which patients self-review an online consent form and provide digital consent). The IRB also approved the present study comparing two IRB-approved methods of MyCode consent using an anonymous survey (IRB# 2017-0334). Patients who agreed to hear about MyCode and completed either IRB-approved consent process were asked if they were willing to complete a brief, anonymous survey about MyCode to inform the MyCode team about the effectiveness of their consent techniques.

Those who provided their verbal agreement were given the survey and included in the analyses. The trial and all survey measures were registered on ClinicalTrials.gov, NCT04131062.

## CODE AVAILABILITY

Access to the R code is available on the project’s OSF page and will be released upon final publication of this paper.

## ACKNOWLEDGEMENTS

The authors thank the MyCode precision health associates who collected data for this study, and the Geisinger patients who participated in the study. This study was funded by the National Cancer Institute of the National Institutes of Health through a bioethics supplement to Award Number R01CA211723 (Alanna Rahm: PI). The content is solely the responsibility of the authors and does not necessarily represent the official views of the National Institutes of Health. The funder played no role in study design, data collection, analysis and interpretation of data, or the writing of this manuscript.

## AUTHOR CONTRIBUTIONS

M.D. and J.W. led the development of the eConsent framework trialed in this study and adapted it for MyCode, with contributions to the latter from J.K.W. and M.N.M. P.R.H., J.W., M.D., J.K.W., and M.N.M. contributed to the conceptualization and methodology of the study, as well as the selection of measures. P.R.H., T.G., R.M., D.R., A.H., J.K.W., and M.N.M. trained MyCode precision health associates on the protocol and oversaw data collection. R.L.V.-Y. led data analyses, with contributions from T.G., R.M., D.R., and A.H. R.L.V.-Y. and M.N.M. wrote the first draft of the paper and all authors edited and reviewed the final manuscript.

## ADDITIONAL INFORMATION

Correspondence should be addressed to Michelle N. Meyer. The views expressed are those of P.R.H. and do not necessarily reflect those of the Consumer Financial Protection Bureau or the United States.

## Footnotes

1 Thirty-one patients were consented while the eConsent application was unavailable (either due to an issue with the iPad, the internet connection, or the app itself). Therefore, these patients could not be randomized (instead, they completed the traditional consent process) and were not enrolled in our consent trial.

2 Answers to the last two questions of the survey (“How often have your health care providers told or given you information about your health and health care that was easy to understand?” and “On a 1-to-5 scale, how would you rate your trust in Geisinger as your health care provider?”) could plausibly have been influenced by the consent intervention. Therefore, we did not calculate the standardized difference between arms for those variables.

3 Comprehension score is significantly associated with other demographic variables in one arm but not the other or overall but not with either arm separately (e.g., with using a personal computer overall or income overall and in the eConsent arm).

4 Note that the power analysis we include here is not post hoc, as we use the same preregistered effect size of interest and did not use any information from the trial data.

